# Mental illness and COVID-19 vaccination: a multinational investigation of observational & register-based data

**DOI:** 10.1101/2024.03.05.24303691

**Authors:** Mary M Barker, Kadri Kõiv, Ingibjörg Magnúsdóttir, Hannah Milbourn, Bin Wang, Xinkai Du, Gillian Murphy, Eva Herweijer, Elísabet U Gísladóttir, Huiqi Li, Anikó Lovik, Anna K. Kähler, Archie Campbell, Maria Feychting, Arna Hauksdóttir, Emily E Joyce, Edda Bjork Thordardottir, Emma M. Frans, Asle Hoffart, Reedik Mägi, Gunnar Tómasson, Kristjana Ásbjörnsdóttir, Jóhanna Jakobsdóttir, Ole A. Andreassen, Patrick F. Sullivan, Sverre Urnes Johnson, Thor Aspelund, Ragnhild Eek Brandlistuen, Helga Ask, Daniel L McCartney, Omid V Ebrahimi, Kelli Lehto, Unnur A Valdimarsdóttir, Fredrik Nyberg, Fang Fang

## Abstract

**Background:** Individuals with mental illness are at higher risk of severe COVID-19 outcomes. However, previous studies on the uptake of COVID-19 vaccination in this population have reported conflicting results. Therefore, we aimed to investigate the association between mental illness and COVID-19 vaccination uptake, using data from five countries.

**Methods:** Data from seven cohort studies (N=325,298), and the Swedish registers (8,080,234), were used to identify mental illness and COVID-19 vaccination uptake. Multivariable modified Poisson regression models were conducted to calculate the prevalence ratio (PR) and 95% CIs of vaccination uptake among individuals with v.s. without mental illness. Results from the cohort studies were pooled using random effects meta-analyses.

**Findings:** Most of the meta-analyses performed using the COVIDMENT study population showed no significant association between mental illness and vaccination uptake. In the Swedish register study population, we observed a very small reduction in the uptake of both the first (prevalence ratio [PR]: 0.98, 95% CI: 0.98-0.99, p<0.001) and second dose among individuals with mental illness; the reduction was however greater among those not using pyschiatric medication (PR: 0.91, 95% CI: 0.91-0.91, p<0.001).

**Conclusions:** The high uptake of COVID-19 vaccination observed among individuals with most types of mental illness highlights the comprehensiveness of the vaccination campaign, however lower levels of vaccination uptake among subgroups of individuals with unmedicated mental illness warrants attention in future vaccination campaigns.

## INTRODUCTION

The coronavirus disease 2019 (COVID-19) pandemic was an unprecedented global health crisis, which, as of August 2023, had caused 6.9 million deaths globally.(1) Although multiple effective vaccines against COVID-19 were developed and distributed globally, vaccine hesitancy and refusal were observed worldwide.(2–5) Crucially, the success of vaccination programmes in controlling the COVID-19 pandemic relies on high vaccination coverage,(6) which is especially important among people at increased risk of severe COVID-19 infection and COVID-19-related mortality, including individuals with mental illness.(7)

Previous systematic reviews exploring the association between mental illness and uptake of various vaccinations have reported heterogenous results.(8, 9) Similarly, results from previous country-specific studies of COVID-19 vaccination uptake in people with mental illness have been inconsistent. While some studies have observed significantly lower uptake of COVID-19 vaccination among individuals with mental illness,(10, 11) others have either reported higher uptake (12), no significant difference,(13) or differences in uptake according to the type of mental illness.(14–16) Additionally, little is currently known regarding the effect of psychiatric treatment on COVID-19 vaccination uptake. As a result, the relationship between mental illness and COVID-19 vaccination uptake may be more complex, and could differ depending on the type, severity, and treatment status of mental illness.

A comprehensive understanding of the impact of mental illness on COVID-19 vaccination uptake is crucial to inform the design of future vaccination campaigns, and, ultimately, to protect at-risk populations from infectious diseases and future pandemics. This study aimed to investigate the association between mental illness and COVID-19 vaccination uptake using data from seven cohort studies included in the multinational COVIDMENT consortium,(17) in addition to data from Swedish national registers, focusing on studying mental illness of different types, severity, and treatment status (i.e., use of prescribed psychiatric medications).

## METHODS

### COVIDMENT Study Analysis

#### Study Population

The COVIDMENT study population included data from seven cohorts within the COVIDMENT consortium, namely the Estonian Biobank (EstBB) cohorts: EstBB full cohort with electronic health record linkages (EstBB-EHR) and the EstBB COVID-19 subcohort (EstBB-C19); the Icelandic COVID-19 National Resilience Cohort (C-19 Resilience); the Norwegian COVID-19 Mental Health and Adherence study (MAP-19); the Norwegian Mother, Father, and Child Cohort study (MoBa);(18) the Scottish CovidLife study; and the Swedish Omtanke2020 study.(17) Ethical approvals for all included cohorts were obtained from regional or national ethics committees, and all participants provided informed consent (Supplementary Table 1). The cohorts used different recruitment strategies, with most recruiting from established cohorts whilst others also allowed self-recruitment via social media. The number of data collection waves varied between the cohorts, with the first data collected in March 2020.

#### Exposure Variables

All exposure variables referred to mental illness experienced before the initiation of COVID-19 vaccination in each study country (Supplementary Table 2). The primary exposure variable was the lifetime diagnosis of any mental illness, which, for all cohorts except EstBB-C19 and EstBB-EHR, was defined using self-report data from questionnaire items asking participants if they had ever been diagnosed with any mental illness. In EstBB-C19 and EstBB-EHR, ICD-10 codes were used to define mental illness diagnoses through linked EHRs (containing diagnoses from Estonian Health Insurance Fund (HIF) treatment bills since 2004), including primary/secondary care and inpatient/outpatient diagnoses (Supplementary Table 3). Anxiety and depressive symptoms were included as secondary exposure variables. In the majority of cohorts, moderate-to-severe anxiety and depressive symptoms, during the past two weeks, were defined as total scores of ≥10 from the Generalised Anxiety Disorder Assessment (GAD-7) and Patient Health Questionnaire-9 (PHQ-9) scales, respectively.(19, 20) In EstBB-C19, anxiety and depressive symptoms, during the past four weeks, were measured using the Emotional State Questionnaire (EST-Q2), utilising cut-off values of >11 to define moderate-to-severe anxiety or depressive symptoms.(21)

#### Outcome Variables

The primary outcome variable was (i) first dose of a COVID-19 vaccine by 30^th^ September 2021. This end date represents the time at which all included countries had offered COVID-19 vaccination to all adult residents and cohort participants had adequate opportunity to book and attend vaccination appointments (Supplementary Table 2). Two additional outcome variables were used: (ii) first dose of a COVID-19 vaccine by 18^th^ February 2022, and (iii) second dose of a COVID-19 vaccine by 18^th^ February 2022. This second date represents the time point at which the majority of cohorts had conducted a new wave of data collection, and therefore it was the first date post-September 2021 that vaccination uptake could be re-assessed. As C-19 Resilience did not conduct further data collection between September 2021 and February 2022, it was only included in the analysis of outcome (i). Participants were eligible for the present study if they had available data for outcomes (i) and/or (ii). Participants were included in the analysis of outcome (iii) if they had received the first dose of any COVID-19 vaccine, except the JCOVDEN vaccine (for which a one-dose schedule was used(22)), before 18^th^ February 2022. In C-19 Resilience, MAP-19, MoBa, and Omtanke2020, vaccination uptake was defined using self-report data collected in various cohort-specific follow-up questionnaires. The remaining cohorts used linked EHR data to define vaccination uptake.

#### Covariates

All covariates (age, sex, smoking status, previous COVID-19 infection, and physical comorbidity status) were defined using data collected before COVID-19 vaccination was initiated in each study country. Smoking was used as a binary variable, defined as current or non-current smoker or user of any tobacco products at the time of data collection. COVID-19 infection was defined as a positive COVID-19 test before the initiation of COVID-19 vaccination. Self-report data was used to define COVID-19 infection in the majority of cohorts. However, the Electronic Communication of Surveillance in Scotland (ECOSS) COVID-19 testing data was used in the CovidLife cohort, and the E-Health Record registry data was used in EstBB-EHR. Physical comorbidity status was defined as the presence of at least two physical health conditions (hypertension, heart disease, lung disease, chronic renal failure, cancer, diabetes, or immunological conditions), using either self-report data from the cohort-specific questionnaires or EHR data (Supplementary Table 3). Neither data on smoking nor physical comorbditiy status was available in the MAP-19 cohort. Further details regarding the time at which variables were defined in each cohort are displayed in Supplementary Table 4.

#### Statistical Analysis

Covariates were summarised, stratified by the diagnosis of any mental illness, using mean (standard deviation [SD]) or frequency (percentage), as appropriate. Multivariable modified Poisson regression models were conducted for each exposure-outcome combination, to assess the prevalence ratio (PR) with 95% confidence intervals (CI), adjusted for age, sex, previous COVID-19 infection, smoking, and physical comorbidity status. Due to data inavailability in MAP-19, models from this cohort were only adjusted for age, sex, and previous COVID-19 infection. As EstBB-EHR had a high proportion of missing COVID-19 testing data, this cohort used an additional “missing” indicator for the COVID-19 infection covariate in addition to “negative” and “positive” groups, to avoid dropping large numbers of participants. Random effects meta-analyses were performed to pool the results from each participating cohort. Heterogeneity was assessed using the I^2^ statistic.

Two sensitivity analyses were conducted: (1) to explore potential differences related to self-report vs. EHR-based variable definitions, meta-analyses for the primary exposure variable (‘diagnosis of any mental illness’) were run excluding cohorts that used EHR to define exposure and/or outcome variables (i.e., CovidLife, EstBB-C19, and EstBB-EHR), (2) to further explore the potential impact of physical health status, meta-analyses for the primary exposure variable were conducted excluding participants with any chronic physical conditions (using all cohorts except MAP-19). Statistical analyses were conducted using STATA (version 17.0) and the metafor package in R (version 4.3.0).(23)

### Swedish Register Study Analysis

#### Study Population

To assess the internal and external validity of the COVIDMENT results, and to gain understanding of the role of disease type, severity and treatment status on the association between mental illness and COVID-19 vaccination uptake, further analyses were conducted using Swedish register data within the SCIFI-PEARL (Swedish COVID-19 Investigation for Future Insights – a Population Epidemiology Approach using Register Linkage) project.(24) Ethical approval was obtained from the Swedish Ethical Review Authority (2020-01800 with subsequent amendments). SCIFI-PEARL is a regularly updated, nationwide register-based study, with individual linkages to multiple registers performed using the unique Swedish Personal Identity Number (PIN). The present study included all individuals aged ≥18 years who were living in Sweden on 27^th^ December 2020 (date of first COVID-19 vaccination in Sweden(25)) and did not die or emigrate on or before the final study end point (30^th^ November 2021).

#### Exposure Variables

Mental illness was first defined using secondary care-based specialist diagnoses (as denoted by ≥1 ICD-10 code listed in Supplementary Table 5) from all inpatient and outpatient hospital encounters reported in the National Patient Register (NPR) between 1^st^ January 2018 and 26^th^ December 2020. In addition to any mental illness, we also studied specific types of mental illness (substance use disorder [excluding aclohool and tobacco use disorders], alcohol use disorder, tobacco use disorder, psychotic disorders, depression, anxiety, and stress-related disorders), defined by ≥1 relevant ICD-10 code. The NPR includes data on specialist care only, whereas patients with milder mental illness are often treated in primary care. Therefore, we also identified information on prescribed use of psychiatric medication (as denoted by ≥1 Anatomical Therapeutic Chemical (ATC) code displayed in Supplementary Table 6), between 1^st^ January 2018 and 26^th^ December 2020, according to the National Prescribed Drug Register (NPDR). Prescribed use of psychiatric medication was used both to ascertain mental illnesses not attended to by specialist care and as a proxy for treatment of mental illness. We studied prescribed use of any psychiatric medication as well as prescribed use of antidepressants, anxiolytics, hypnotics/sedatives, and antipsychotics, respectively.

To investigate the effect of disease severity and treatment with psychiatric medication, an alternative (multi-level) categorisation of exposure variables was used for any mental illness, depression, anxiety, and psychotic disorder, with the following categories: (1) no specialist diagnosis of mental illness and no prescribed use of psychiatric medication (i.e., reference category), (2) prescribed use of psychiatric medication without specialist diagnosis (i.e., proxy of milder illness with medical treatment), (3) specialist diagnosis with prescribed use of psychiatric medication (i.e., proxy of more severe illness with medical treatment), and (4) specialist diagnosis without prescribed use of psychiatric medication (i.e., proxy of more severe illness without medical treatment).

#### Outcome Variables

For the definition of outcome variables, the National Vaccination Register (NVR) was used, which includes information on all COVID-19 vaccinations conducted in Sweden from December 2020.(24) Two outcome variables were used: (i) first dose of a COVID-19 vaccine by 30^th^ September 2021, and (ii) second dose of a COVID-19 vaccine by 30^th^ November 2021. By using this end date for outcome (ii) we ensured that all individuals vaccinated with a first dose up until 30^th^ September would have had the recommended time period between their first and second vaccine dose.(26) Individuals were included in the analysis of outcome (ii) if they had received the first dose of any COVID-19 vaccine, except the JCOVDEN vaccine, by 30^th^ September 2021.

#### Covariates

Covariates included age and sex (identified through the Total Population Register (TPR)), region of residence, highest educational attainment, cohabitation status, and income (identified from the Swedish Longitudinal Integrated Database for Health Insurance and Labour Market Studies (LISA) in 2020), severe COVID-19 infection and the Charlson Comorbidity Index (CCI). Severe COVID-19 infection was defined as an inpatient or ICU visit due to COVID-19 before the study start date (27^th^ December 2020), whilst CCI was calculated at the study start date, using diagnostic data identified from 1^st^ January 2015 onwards in the NPR.(27)

#### Statistical Analysis

All covariates were summarized, stratified by the diagnosis of any mental illness, using mean (standard deviation [SD]) or frequency (percentage), as appropriate. Multivariable modified Poisson regression models were run for all exposures and outcomes to assesss PR and 95% CIs of COVID-19 vaccination uptake in relation to mental illness, adjusted for the covariates listed above. Stratified analyses were conducted by sex and the presence of chronic physical condition(s) (defined as a CCI score of ≥1). Complete case analysis was used throughout. Statistical analyses were conducted using STATA (version 17.0).

## RESULTS

### COVIDMENT Study Analysis

Of the 403,794 individuals included in the participating COVIDMENT cohort studies, 325,298 individuals met the eligibility criteria for the present study (Supplementary Figure 1). Over half of the overall study population (65.1%), and of each participating cohort, were female (Supplementary Table 7). The mean age in the participating cohorts ranged from 36.9 years (MAP-19) to 59.4 years (CovidLife), with a mean of 48 years in the overall study population.

Individuals with a diagnosis of any mental illness were more likely to be female, compared to those without a mental illness (Table 1). The proportion of individuals with chronic physical conditions was higher among those with any mental illness diagnosis. Low levels of missing data were observed for the majority of covariates.

**Table 1:** Distribution of sociodemographic variables in the included COVIDMENT study population, overall and by diagnosis of any mental illness diagnosis, presented as N (%) or mean [SD].

|  | Diagnosis of any mental illness |  |  |  |
| --- | --- | --- | --- | --- |
|  | Yes<br>(n=122,976) | No (n=201,273) | Missing<br>(n=1,049) | Total<br>(N=325,298) |
| Cohort |  |  |  |  |
| EstBB-C19 (Estonia) | 3,107 (2.5%) | 2,526 (1.3%) | 0 (0.0%) | 5,633 (1.7%) |
| EstBB-EHR (Estonia) | 95,208 (77.4%) | 88,124 (43.8%) | 0 (0.0%) | 183,332 (56.4%) |
| C-19 Resilience (Iceland) | 2,895 (2.4%) | 7,283 (3.6%) | 239 (22.8%) | 10,417 (3.2%) |
| MAP-19 (Norway) | 698 (0.6%) | 2,722 (1.3%) | 474 (45.2%) | 3,894 (1.2%) |
| MoBa (Norway) | 15,496 (12.6%) | 87,315 (43.4%) | 0 (0.0%) | 102,811 (31.6%) |
| CovidLife (Scotland) | 1,230 (1.0%) | 3,499 (1.7%) | 31 (2.9%) | 4,760 (1.5%) |
| Omtanke2020 (Sweden) | 4,342 (3.5%) | 9,804 (4.9%) | 305 (29.1%) | 14,451 (4.4%) |
| Sex |  |  |  |  |
| Female | 88,515 (72.0%) | 122,544 (60.9%) | 753 (71.8%) | 211,812 (65.1%) |
| Male | 34,446 (28.0%) | 78,713 (39.1%) | 205 (19.5%) | 113,364 (34.9%) |
| Other | 8 (0.0%) | 6 (0.0%) | 1 (0.1%) | 15 (0.0%) |
| Missing | 7 (0.0%) | 10 (0.0%) | 90 (8.6%) | 107 (0.0%) |
| Age group, years |  |  |  |  |
| 18-29 | 12,826 (10.5%) | 14,670 (7.3%) | 265 (25.3%) | 27,761 (8.5%) |
| 30-39 | 24,119 (19.6%) | 30,486 (15.1%) | 197 (18.8%) | 54,802 (16.9%) |
| 40-49 | 31,864 (25.9%) | 78,311 (38.9%) | 177 (16.9%) | 110,352 (33.9%) |
| 50-59 | 24,359 (19.8%) | 44,785 (22.2%) | 184 (17.5%) | 69,328 (21.3%) |
| 60-69 | 17,228 (14.0%) | 19,053 (9.5%) | 147 (14.0%) | 36,428 (11.2%) |
| 70+ | 12,558 (10.2%) | 13,845 (6.9%) | 76 (7.2%) | 26,479 (8.1%) |
| Missing | 22 (0.0%) | 123 (0.1%) | 3 (0.3%) | 148 (0.1%) |
| Mean [SD] age, years | 48.5 [1.8] | 47.8 [3.6] | 44.1 [9.4] | 48.0 [2.9] |
| COVID-19 infection |  |  |  |  |
| Yes | 3,094 (2.5%) | 4,629 (2.3%) | 17 (1.6%) | 7,740 (2.4%) |
| No | 64,035 (52.1%) | 140,877 (70.0%) | 1032 (98.4%) | 205,944 (63.3%) |
| Missing* | 55,847 (45.4%) | 55,767 (27.7%) | 0 (0.0%) | 111,614 (34.3%) |
| Smoking status |  |  |  |  |
| Yes | 26,465 (21.5%) | 34,253 (17.0%) | 62 (5.9%) | 60,780 (18.7%) |
| No | 86,737 (70.5%) | 155,181 (77.1%) | 493 (47.0%) | 242,411 (74.5%) |
| Missing | 9,774 (8.0%) | 11,839 (5.9%) | 494 (47.1%) | 22,107 (6.8%) |
| Chronic physical conditions |  |  |  |  |
| 0 | 47,606 (38.7%) | 123,948 (61.6%) | 160 (15.2%) | 171,714 (52.8%) |
| 1 | 34,723 (28.2%) | 46,836 (23.3%) | 68 (6.5%) | 81,627 (25.1%) |
| 2+ | 39,497 (32.1%) | 27,288 (13.5%) | 46 (4.4%) | 66,831 (20.5%) |
| Missing | 1,150 (0.9%) | 3,201 (1.6%) | 775 (73.9%) | 5,126 (1.6%) |

\*Mainly due to COVID-19 testing data only being available for some EstBB-EHR individuals

#### First dose of a COVID-19 vaccine by 30^th^ September 2021

314,827 individuals were included in the analysis of uptake of the first dose of a COVID-19 vaccine by 30^th^ September 2021 (Table 2). Overall vaccination uptake was high (85.1%; n=267,981/314,827). However, a small difference in uptake was observed between individuals with (82.4%; n=99,041/120,212) v.s. without (86.8%; n=168,174/193,706) any mental illness. Vaccination uptake in each included cohort is displayed in Supplementary Table 8.

**Table 2:** Uptake of COVID-19 vaccination overall and by diagnosis of any mental illness diagnosis, in the included COVIDMENT study population, presented as N (%)

|  | Uptake of first dose of a COVID-19 vaccine by 30<sup>th</sup> September 2021 |  |  | Uptake of first dose of a COVID-19 vaccine by 18<sup>th</sup> February 2022 |  |  | Uptake of second dose of a COVID-19 vaccine by 18<sup>th</sup> February 2022 |  |  |
| --- | --- | --- | --- | --- | --- | --- | --- | --- | --- |
|  | Yes | No | Total | Yes | No | Total | Yes | No | Total |
| Total study population | 267,981<br>(85.1%) | 46,846<br>(14.9%) | 314,827<br>(100.0) | 278,887<br>(88.9%) | 34,697<br>(11.1%) | 313,584<br>(100.0%) | 252,439<br>(95.5%) | 11,965<br>(4.5%) | 264,404<br>(100.0%) |
| Mental illness | 99,041<br>(82.4%) | 21,171<br>(17.6%) | 120,212<br>(100.0%) | 103,955<br>(86.7%) | 15,953<br>(13.3%) | 119,908<br>(100.0%) | 93,420<br>(94.7%) | 5,251<br>(5.3%) | 98,671<br>(100.0%) |
| No mental illness | 168,174<br>(86.8%) | 25,532<br>(13.2%) | 193,706<br>(100.0%) | 174,612<br>(90.3%) | 18,728<br>(9.7%) | 193,340<br>(100.0%) | 158,830<br>(95.9%) | 6,712<br>(4.1%) | 165,542<br>(100.0%) |
| Missing | 766<br>(84.3%) | 143<br>(15.7%) | 909<br>(100.0%) | 320<br>(95.2%) | 16 (4.8%) | 336<br>(100.0%) | 189<br>(99.0%) | 2 (1.0%) | 191<br>(100.0%) |

Results from the meta-analysis showed no significant association, after adjustment for covariates, between the diagnosis of any mental illness and vaccination uptake (pooled PR: 0.99, 95% CI: 0.97-1.00]; I^2^: 91.7%, p<0.001) (Figure 1A, Supplementary Table 9). Although the level of heterogeneity was high, a statistically significant association between the diagnosis of any mental illness and lower vaccination uptake was only found in the EstBB-EHR and MAP-19 cohorts. No associations were observed between anxiety or depressive symptoms and vaccination uptake.

**Figure 1:**
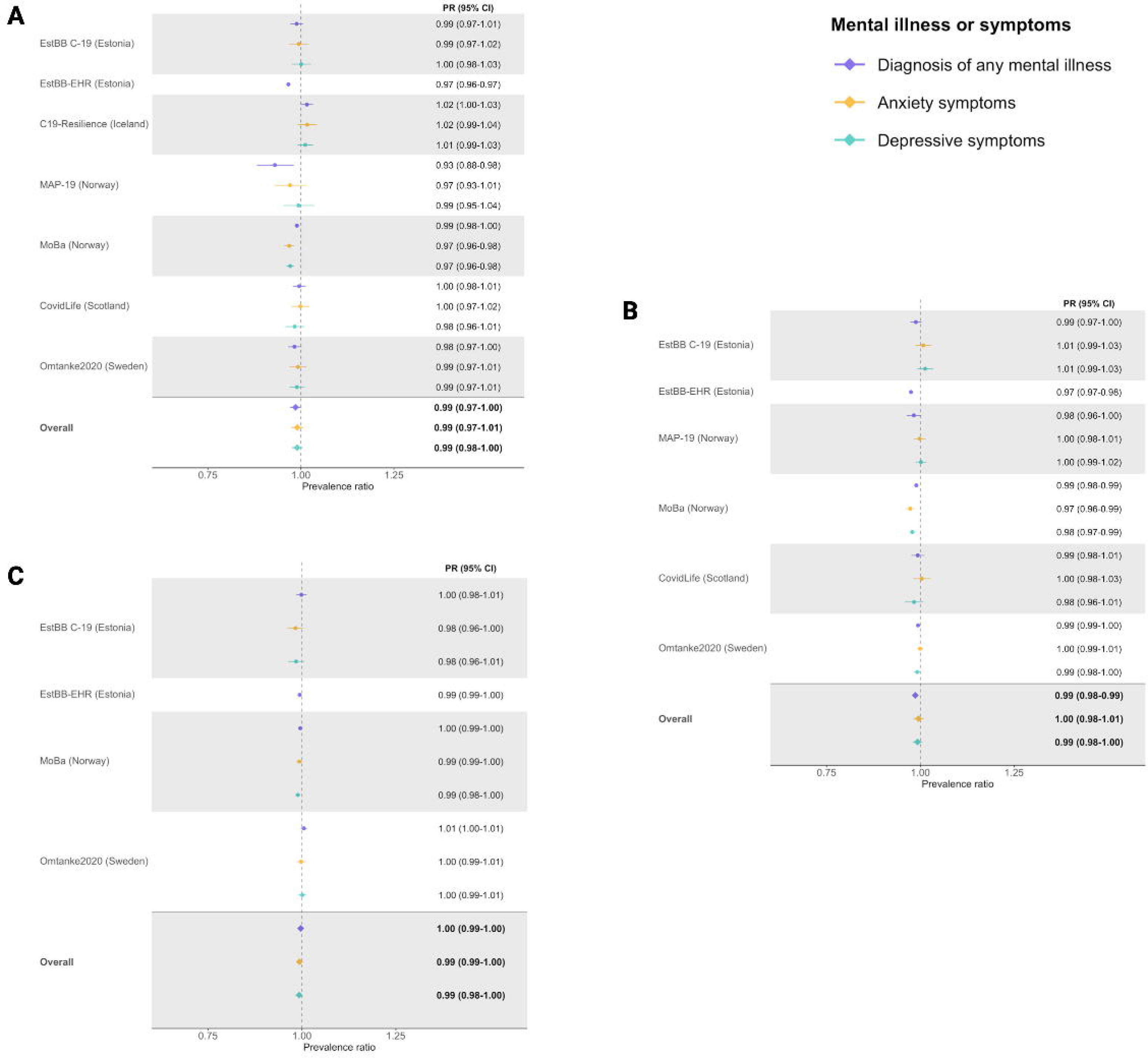
Prevalence ratio (PR) and 95% CI of (A) first dose of a COVID-19 vaccine by 30^th^ September 2021, (B) first dose of a COVID-19 vaccine by 18^th^ February 2022, (C) second dose of a COVID-19 vaccine by 18^th^ February 2022, according to the presence of any mental illness diagnosis, anxiety symptoms or depressive symptoms, in the included COVIDMENT study population. N.B. PR and 95% CI reported rounded to 2 decimal places.

#### First dose of a COVID-19 vaccine by 18^th^ February 2022

Uptake of the first dose of a COVID-19 vaccine by 18^th^ February 2022 was analysed in 313,584 individuals. Vaccination uptake was high (88.9%; n=278,887/313,584); however, a small difference in uptake remained between individuals with (86.7%; n=103,955/119,908) v.s. without (90.3%; n=174,612/193,340) mental illness (Table 2).

Results from the meta-analysis revealed a small association, after adjustment for covariates, between the diagnosis of any mental illness and vaccination uptake (PR: 0.99, 95% CI: 0.98-0.99; I^2^: 79.98, p<0.001) (Figure 1B, Supplementary Table 9). No associations were observed between anxiety or depressive symptoms and vaccination uptake.

#### Second dose of a COVID-19 vaccine by 18^th^ February 2022

264,404 individuals were eligible for the analysis of uptake of the second dose of a COVID-19 vaccine by 18^th^ February 2022 (Table 2). Among these individuals, vaccination uptake wasvery high (95.5%; n=252,439/264,404) and the difference in uptake between those with (94.7%; n=93,420/98,671) v.s. without (95.9%; n=158,830/165,542) mental illness was very small. Due to low numbers of participants, models could not be run in the MAP-19 and CovidLife cohorts.

The meta-analysis of the remaining eligible cohorts showed no significant differences in vaccination uptake by the diagnosis of any mental illness or the presence of anxiety or depressive symptoms, after adjustment for covariates (Figure 1C, Supplementary Table 9).

For all outcomes, no substantial differences were observed in any of the sensitivity analyses (Supplementary Table 10).

### Swedish Register Study Analysis

Among the 8,080,234 individuals included in the Swedish register study population, individuals with a specialist diagnosis of mental illness were more likely to be female (55.5% vs. 49.7%) and were, on average, younger (45.0 years vs. 50.2 years), compared to those without a mental illness (Table 3). Among those with a mental illness diagnosis, the proportions of individuals who had completed university education, were cohabiting, and were in the highest quartile of income were all lower, whilst the presence of chronic physical conditions was more common, compared to those without a mental illness diagnosis. The proportion of missing data for all covariates was low (≤2.5%).

**Table 3:**
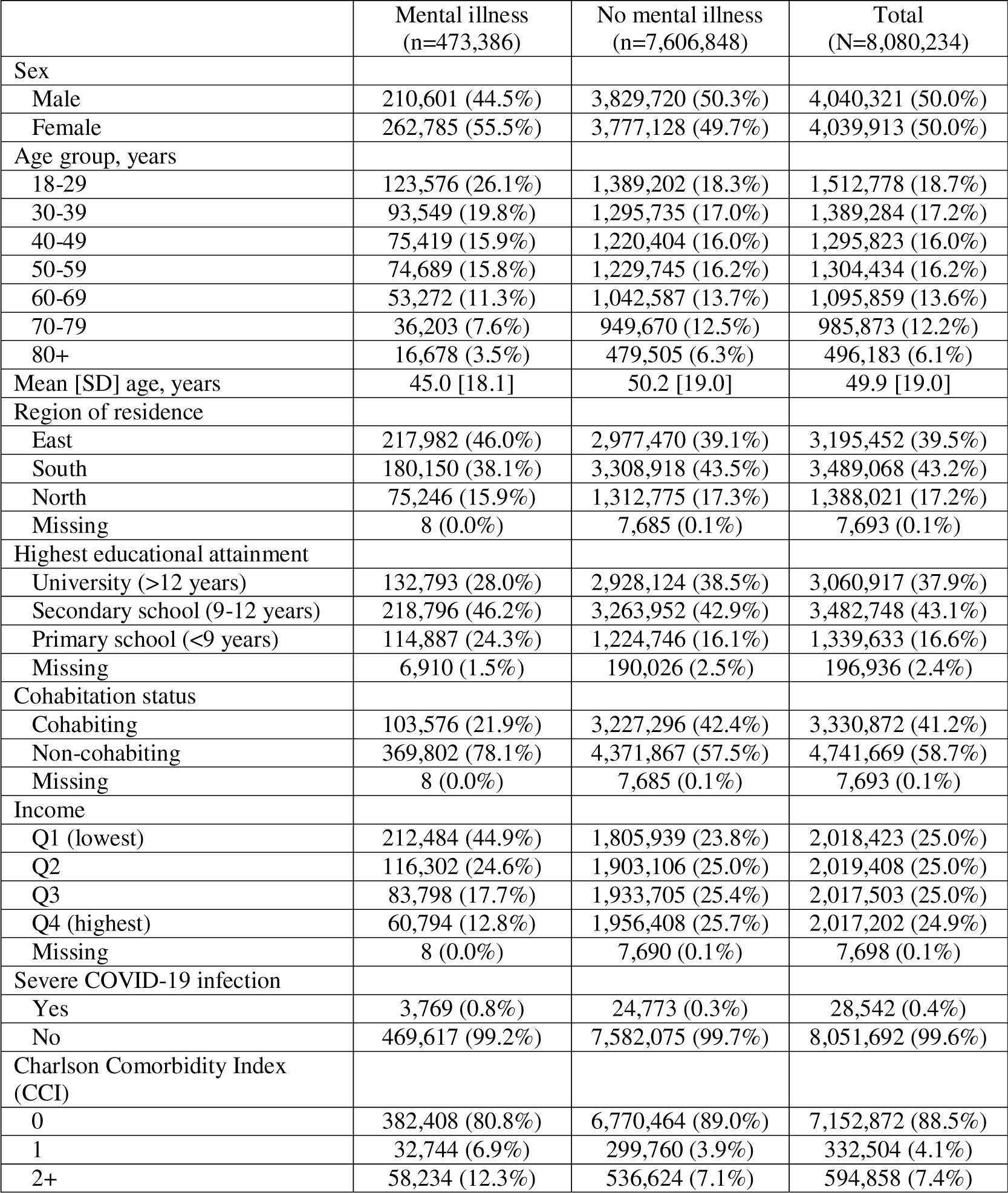
Distribution of sociodemographic variables in the included Swedish register study population, overall and by specialist diagnosis of a mental illness, presented as N (%) or mean [SD].

#### First dose of a COVID-19 vaccine by 30^th^ September 2021

In this study population, uptake of the first dose of a COVID-19 vaccine by 30^th^ September 2021 was high (84.6%; n=6,834,074/8,080,234) (Table 4). However vaccination uptake was slightly lower in individuals with (78.1%; n=369,549/473,386) v.s. without (85.0%; 6,464,525/7,606,848) a specialist diagnosis of a mental illness. Vaccination uptake in relation to each type of mental illness diagnosis and type of psychiatric medication used is shown in Supplementary Table 11.

**Table 4:** Uptake of COVID-19 vaccination overall, and by specialist diagnosis of any mental illness and prescribed use of any psychiatric medication, in the included Swedish register population, presented as N (%)

|  | First dose of a COVID-19 vaccine by 30<sup>th</sup> September 2021 |  |  | Second dose of a COVID-19 vaccine by 30<sup>th</sup> November 2021 |  |  |
| --- | --- | --- | --- | --- | --- | --- |
|  | Yes | No | Total | Yes | No | Total |
| Total study population | 6,834,074<br>(84.6%) | 1,246,160 (15.4%) | 8,080,234 (100.0%) | 6,704,293<br>(98.1%) | 129,761 (1.9%) | 6,834,054 (100.0%) |
| Any mental illness |  |  |  |  |  |  |
| Yes | 369,549 (78.1%) | 103,837 (21.9%) | 473,386 (100.0%) | 355,040 (96.1%) | 14,508 (3.9%) | 369,548 (100.0%) |
| No | 6,464,525<br>(85.0%) | 1,142,323 (15.0%) | 7,606,848 (100.0%) | 6,349,253<br>(98.2%) | 115,253 (1.8%) | 6,464,506 (100.0%) |
| Any psychiatric medication |  |  |  |  |  |  |
| Yes | 1,801,401<br>(87.7%) | 253,406 (12.3%) | 2,054,807 (100.0%) | 1,766,489<br>(98.1%) | 34,909 (1.9%) | 1,801,398 (100.0%) |
| No | 5,032,673<br>(83.5%) | 992,754 (16.5%) | 6,025,427 (100.0%) | 4,937,804<br>(98.1%) | 94,852 (1.9%) | 5,032,656 (100.0%) |

Taking into account all covariates, we found that the uptake of vaccination in individuals with any mental illness was 2% lower than that of individuals without a mental illness (PR: 0.98, 95% CI: 0.98-0.99, p<0.001) (Figure 2A). Similarly small differences in vaccination uptake were shown for most types of mental illness, except for substance use disorder which had the strongest association with lower vaccination uptake (PR: 0.95, 95% CI: 0.94-0.95, p<0.001). A 3% higher uptake was observed among individuals using prescribed psychiatric medication, compared to individuals not using such medication (PR: 1.03, 95% CI: 1.03-1.03, p<0.001). Similar associations were found for different types of psychiatric medication. Stratified analyses revealed no substantial differences in the associations by sex or the presence of chronic physical conditions (Supplementary Table 12).

**Figure 2:**
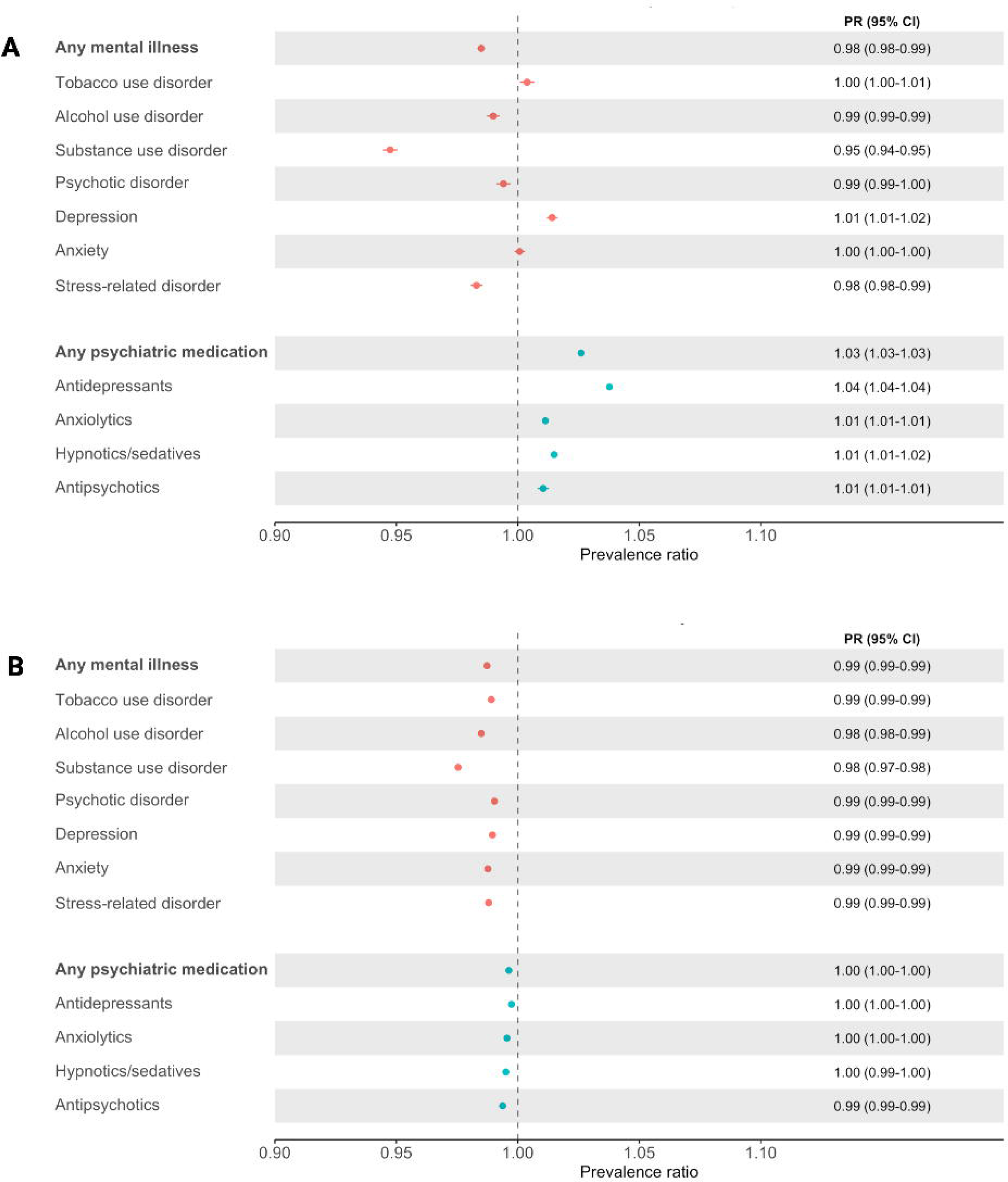
Prevalence ratio (PR) and 95% CI of (A) first dose of a COVID-19 vaccine by 30^th^ September 2021 and (B) second dose of a COVID-19 vaccine by 30^th^ November 2021, according to specialist diagnosis of a mental illness and prescribed use of psychiatric medication, in the Swedish register study population. N.B. PR and 95% CI reported rounded to 2 decimal places. ‘Substance use disorder’ excludes alcohol and tobacco use disorders.

Results from the multi-level exposure analysis showed that, compared to individuals with neither a specialist mental illness diagnosis nor prescribed use of psychiatric medication, vaccination uptake was 3% higher among those with prescribed use of psychiatric medication but no specialist diagnosis (PR: 1.03, 95% CI: 1.03-1.03, p<0.001), and 1% higher among those with both a specialist diagnosis and prescribed use of any psychiatric (PR: 1.01, 95% CI: 1.01-1.01, p<0.001) (Figure 3A). However, those with a specialist diagnosis of any mental illness but no prescribed use of any psychiatric medication had a 9% reduction in vaccination uptake (PR: 0.91, 95% CI: 0.91-0.91, p<0.001). This pattern was also observed for the multi-level exposure analysis carried out for anxiety, depression, and psychotic disorder.

**Figure 3:**
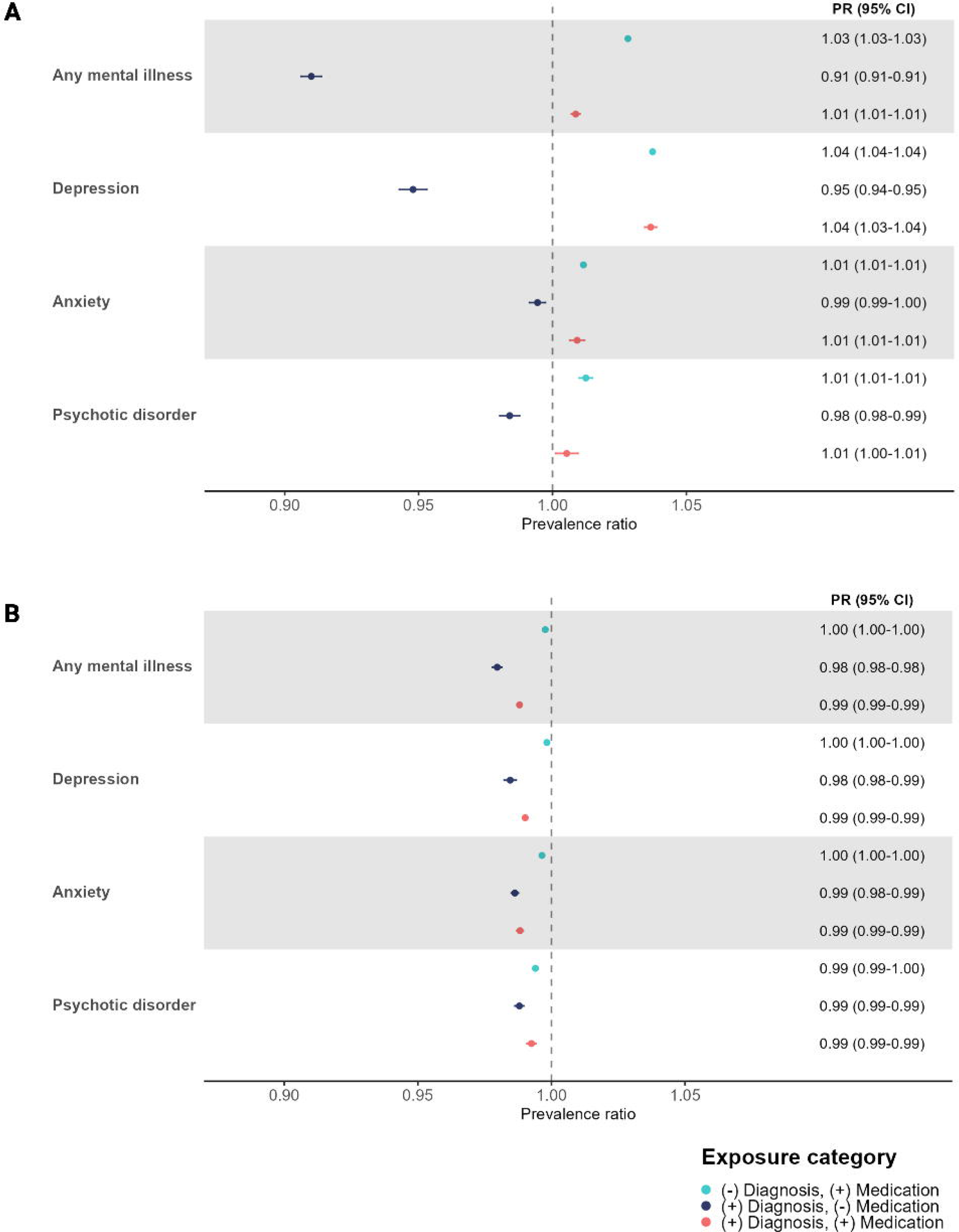
Prevalence ratio (PR) and 95% CI of (A) first dose of a COVID-19 vaccine by 30^th^ September 2021 and (B) second dose of a COVID-19 vaccine by 30^th^ November 2021, according to specialist diagnosis of a mental illness/prescribed medication use status, in the Swedish register study population. N.B. PR and 95% CI reported rounded to 2 decimal places.

#### Second dose of a COVID-19 vaccine by 30^th^ November 2021

6,834,054 individuals were eligible for the analysis of the uptake of the second dose of a COVID-19 vaccine by 30^th^ November 2021 (Table 4). Vaccination uptake had a very high coverage overall (98.1%; n=6,704,293/6,834,054), and the difference between those with (96.1%; n=355,040/369,548) v.s. without (98.2%; n=6,349,253/6,464,506) a mental illness was very small.

A very small difference in vaccination uptake was noted among those with v.s. without a specialist diagnosis of a mental illness (PR: 0.99, 95% CI: 0.99-0.99, p<0.001) or prescribed use of psychiatric medication (Figure 2B). This was also observed for different types of mental illness diagnosis and all types of psychiatric medication. Results from the stratified analyses showed no differences in the associations by sex or the presence of chronic physical conditions (Supplementary Table 12). Simiarly, the multi-level exposure analysis showed statistically significant, but very small, differences in vaccination uptake according to specialist diagnosis and/or prescribed medication use (Figure 3B).

## DISCUSSION

This multinational study of 325,298 individuals from the participating COVIDMENT cohort studies, and 8,080,234 individuals from the Swedish national registers, showed that uptake of COVID-19 vaccination was high, and differences in uptake by mental illness were, in general, small. The majority of the analyses conducted in the COVIDMENT study population showed no significant difference in vaccination uptake according to the presence of diagnosed mental illness or anxiety or depressive symptoms. In the Swedish register analysis, although slightly lower vaccination uptake was observed among individuals with a specialist diagnosis of a mental illness, the absolute difference in vaccination uptake was very small. We did, however, observe that individuals with a specialist diagnosis of mental illness without ongoing psychiatric treatment had approximately 9% lower uptake of COVID-19 vaccination.

Our findings of substantially lower vaccination uptake among individuals with unmedicated diagnosed mental illness in the Swedish register study analysis have important implications. Although we were unable to investigate the underlying reasons in the present study, lower vaccination uptake could have been due to particularly low levels of engagement with preventative healthcare in these groups.(28) Individuals with mental illness have been shown to have poor access to nonpsychiatric healthcare, including preventative services such as vaccination programmes, primarily due to barriers such as low levels of knowledge and awareness of such services, and accessibility issues.(29) However, pilot interventions aimed at increasing vaccination uptake among individuals with mental illness by addressing these barriers, for example through targeted education campaigns and the integration of psychiatric providers in vaccination programmes, have been shown to be effective.(29, 30) Therefore, strategies such as these could be incorporated into future vaccination campaings in order to reduce barriers to vaccination among individuals with relatively severe mental illness (e.g., attended by specialist care).

We found no significant association between mental illness and COVID-19 vaccinion uptake in most of the COVIDMENT cohorts, and the absolute rate of vaccination in the included cohorts was generally high, both among individuals with and without mental illness. The slight differences across cohorts could have been, at least partially, due to the varying prioritisation schedules for COVID-19 vaccination used in the different countries. Although the European Union (EU) recommended that member states prioritise indivdiuals at the highest risk for severe COVID-19, countries could decide which population groups to include in their prioritisation schedules, meaning that not all countries prioritised vaccination for individuals with mental illness.(31)

There are many strengths of our study, which represents the first multinational study of the association between mental illness and COVID-19 vaccination uptake. The complementary use of the two different types of prospectively collected data sources combined the benefits of the rich self-reported COVIDMENT data with the Swedish national registers, which has minimal concerns of selection bias, thereby increasing the robustness of the study’s conclusions. By including a very large sample size from several different countries we were also able to investigate whether the results found in previous country-specific studies translated to an multinational context.

However, limitations of the study must also be noted. Although the multinational nature of the study increases the representativeness of the findings, all participating countries have established welfare systems and generally accessible healthcare, meaning caution should be taken when generalising the results to other global regions. There were also specific limitations relating to the two study populations, such as potential selection bias in the COVIDMENT cohorts, meaning individuals with severe mental illness were less likely to take part in the study.

In conclusion, in this large, multinational study we showed that uptake of the COVID-19 vaccine was high, even among most individuals with a mental illness, highlighting the comprehensiveness and success of the COVID-19 vaccination campaign in reaching most population groups. However, specific groups of people with a recent specialist diagnosis of mental illness yet not on psychiatric medication, were still at risk for low vaccination uptake. These findings have important implications for the design of current and future vaccination campaigns against infectious diseases and future pandemics.

## Data Availability

The raw data underlying this article were subject to ethical approval and cannot be shared publicly due to data protection laws in each participating country.

## Acknowledgements

We would like to thank all of the participants who contributed data to the COVIDMENT project. The Norwegian Mother, Father and Child Cohort Study is supported by the Norwegian Ministry of Health and Care Services and the Ministry of Education and Research. We are grateful to all the participating families in Norway who take part in this ongoing cohort study. We would also like to acknowledge the work of the Estonian Biobank Research Team.

