## Supplementary Table 1 for "Mental illness and COVID-19 vaccination: a multinational investigation of observational & register-based data"

[Supplementary Table 7: Distribution of sociodemographic variables in the total included COVIDMENT study population, and in each participating cohort, presented as N (%) or mean [SD].	8](#_Toc152575387)

[Supplementary Table 10: Sensitivity model results (pooled prevalence ratio [PR] (95% CI)), according to the presence of any mental illness diagnosis, in the included COVIDMENT study population.	12](#_Toc152575390)

### **Supplementary Table 1:** Ethical approvals obtained for each included COVIDMENT cohort study.

| Cohort | Ethical Approval |
| --- | --- |
| EstBB cohorts | Estonian Committee on Bioethics and Human Research (1.1–12/1277 and 1.1–12/2860) |
| C-19 Resilience | National Bioethics Committee (NBC no. 20– 073, 21–071) as well as the National Data Protection Authority |
| MAP-19 | Regional Committee for Medical Research Ethics, reference number: 125510 |
| MoBa | Regional Committees for Medical and Health Research Ethics (127708/14140/20138) |
| CovidLife | East of Scotland Research Ethics Service (EoSRES) |
| Omtanke2020 | Ethical approval no. 2020–01785 |

EstBB cohorts (EstBB-C19 = The Estonian Biobank COVID-19 Cohort; EstBB-EHR = The Estonian Biobank electronic health records); C19-Resilience = The Icelandic COVID-19 National Resilience Cohort; MAP-19 = The Norwegian COVID-19, Mental Health and Adherence Project; MoBa = The Norwegian Mother, Father and Child Cohort Study

### **Supplementary Table 2:** Key COVID-19 vaccination dates for countries included in the COVIDMENT study population.

| Country | Date of first COVID-19 vaccination | Date at which all adults offered at least one dose of a COVID-19 vaccine |
| --- | --- | --- |
| Estonia | January 2021^1^ | May 2021^2^ |
| Iceland | December 2020^3^ | June 2021^4^ |
| Norway | December 2020^5^ | March 2021^6^ |
| Scotland | December 2020^7^ | June 2021^8^ |
| Sweden | 27th December 2020^9^ | August- September 2021*^10-12^ |

*dependent on region of residence

^8^ Scottish Government. First doses booked in for all adults ahead of schedule. 2021. Accessed: 10 October 2022. Available from:

https://www.gov.scot/news/first-doses-booked-in-for-all-adults-ahead-of-schedule/

^9^ Krisinformation.se. Covid-19 vaccination begins on 27 December. 2020 10 October 2022]; Available from: https://www.krisinformation.se/en/news/2020/december/vaccination-27-december

^10^ Sveriges Kommuner och Regioner [SKR]. Regionernas planering avseende vaccinering mot covid-19, delrapport 6 [The planning regarding vaccination against Covid-19 for regions, report 6]. 2021. Accessed: 10 October 2021. Available from: https://skr.se/download/18.5bb54e0c179a302981232fd/1621980314035/Regionernas_planer%20ing_%20vaccinering_covid-19_delrapport%206.pdf

^11^ Krisinformation.se. Vecka 26 2021. Accessed: 10 October 2022. 2021. Available from: https://www.krisinformation.se/om-krisinformation/for-myndigheter-och-andra-aktorer/omvarldsbevakning/20212/vecka-26-2021

^12^ Folkhälsomyndigheten. Över 70 procent av Sveriges vuxna befolkning har fått vaccin mot covid-19. 2021. Accessed: 10 October 2022. Available from: <https://www.folkhalsomyndigheten.se/nyheter-och-press/nyhetsarkiv/2021/juli/over-70-procent-av-sveriges-vuxna-befolkning-har-fatt-vaccin-mot-covid-19/>

### **Supplementary Table 3:** ICD-10 codes used to define mental illness (in the EstBB-C19 and EstBB-EHR cohorts) and physical comorbidity status (in the EstBB-EHR cohort).

| Variable | Condition | ICD-10 codes |
| --- | --- | --- |
| Any mental illness | | F00-F99 |
| Physical comorbidity status | Hypertension | I10 |
|  | Heart disease | I00-I99 (excluding I10) |
|  | Lung disease | J12-J18, J21-J22, J40-J47, J60-J70, J80-J84, E84.0, Q26.8, Q89.3, M05.1 |
|  | Chronic renal failure | N17-N19 |
|  | Cancer | C00-C97 |
|  | Diabetes | E10-E14 |
|  | Immunological conditions | B17-B40, D69.0, D69.3, D80-D89, E05.0, E06.3, E10-E14, G04.0, G04.8, G05.8, G35, G36.0-G36.1, G36.8-G36.9, G37.2-G37.5, G37.8-G37.9,G61.0, G70.0, H46.9, L10.0, L10.2, L12.0-L12.3, L12.8-L12.9, M05.0-M05.3, M05.8-M05.9, M06.0-M06.4, M06.8-M06.9, M08.0, M08.2-M08.4, M08.8-M08.9, M31.0, M32, M35.0, N08.5 |

### **Supplementary Table 4:** Timing of variable definitions in the participating COVIDMENT cohorts.

| Variable | Cohort | Timing of variable measurement |
| --- | --- | --- |
| Exposure variables | | |
| Diagnosis of any mental illness | EstBB-C19 | Diagnosis from HIF bills (2004 - 26^th^ December 2020) |
|  | EstBB-EHR | Diagnosis from HIF bills (2004 - 26^th^ December 2020) |
|  | C-19 Resilience | Baseline questionnaire (24^th^ April 2020-29^th^ December 2020) |
|  | MAP-19 | Participants asked about mental illness diagnosis prior to the start of the COVID-19 pandemic in the following follow-up questionnaires: 24^th^ October-12^th^ November 2021, 2^nd^-14^th^ January 2022, 6^th^-27^th^ March 2022 |
|  | MoBa | For females: 1999-2008, 2007-2017; for males: 1999-2008, 2015  Responses were collected from the first questionnaire collected at recruitment (during pregnancy) between 1999-2009. Additional cases were collected from self-reported mental illnesses in questionnaires responded to when the children were 8 years old (between 2007 and 2017) for females (mothers), and in 2015 for males (fathers).  Participants could answer ”yes” (defined as mental illness diagnosis) or give no response. |
|  | CovidLife | Baseline questionnaire (COVIDLife1: 17th April -7th June 2020) |
|  | Omtanke2020 | Baseline questionnaire (9^th^ June 2020-26th December 2020) |
| Anxiety symptoms | EstBB-C19 | C-19 questionnaire (10^th^ May 2020 – 26^th^ December 2020) |
|  | EstBB-EHR | NA |
|  | C-19 Resilience | Baseline questionnaire and monthly follow-up questionnaires (24^th^ April 2020-29^th^ December 2020) |
|  | MAP-19 | Baseline questionnaire (31st March-7th April 2020) |
|  | MoBa | Biweekly follow-up questionnaires (11^th^ May 2020-24^th^ May 2020) |
|  | CovidLife | Baseline questionnaire (COVIDLife1: 17th April -7th June 2020), second questionnaire (COVIDLife2: 21st July - 16th August 2020) |
|  | Omtanke2020 | Baseline and monthly follow-up questionnaires (9^th^ June 2020-26^th^ December 2020) |
| Depressive symptoms | EstBB-C19 | C-19 questionnaire (10^th^ May 2020 – 26^th^ December 2020) |
|  | EstBB-EHR | NA |
|  | C-19 Resilience | Baseline questionnaire and monthly follow-up questionnaires (24^th^ April 2020-29^th^ December 2020) |
|  | MAP-19 | Baseline questionnaire (31st March-7th April 2020) |
|  | MoBa | Biweekly follow-up questionnaires (10^th^ June 2020-23^rd^ June 2020) |
|  | CovidLife | Baseline questionnaire (COVIDLife1: 17th April -7th June 2020), second questionnaire (COVIDLife2: 21st July - 16th August 2020) |
|  | Omtanke2020 | Baseline and monthly follow-up questionnaires (9^th^ June 2020-26^th^ December 2020) |
| Outcome variables | | |
| First dose of a COVID-19 vaccine by 30^th^ September 2021 | EstBB-C19 | e-Health Record registry immunization notices |
|  | EstBB-EHR | e-Health Record registry immunization notices |
|  | C-19 Resilience | Monthly follow-up questionnaires (27^th^ May 2021-28^th^ August 2021) |
|  | MAP-19 | Follow-up questionnaire (4^th^ July-1^st^ August 2021) |
|  | MoBa | Biweekly follow-up questionnaires (2^nd^ February 2021-30^th^ September 2021) |
|  | CovidLife | Scotland National COVID-19 vaccination data (8^th^ December 2020 – 30^th^ September 2021) |
|  | Omtanke2020 | Monthly follow-up questionnaires (27^th^ Dec 2020-30^th^ September 2021) |
| First dose of a COVID-19 vaccine by 18^th^ February 2022 | EstBB-C19 | e-Health Record registry immunization notices |
|  | EstBB-EHR | e-Health Record registry immunization notices |
|  | C-19 Resilience | NA |
|  | MAP-19 | Follow-up questionnaire (2^nd^-14^th^ January 2022) |
|  | MoBa | Biweekly follow-up questionnaires (2^nd^ February 2021-18^th^ February 2022) |
|  | CovidLife | Scotland National COVID-19 vaccination data (8^th^ December 2020 – 30^th^ September 2021) |
|  | Omtanke2020 | Annual follow-up questionnaire (1^st^ December 2021-18^th^ February 2022) |
| Second dose of a COVID-19 vaccine by 18^th^ February 2022 | EstBB-C19 | e-Health Record registry immunization notices |
|  | EstBB-EHR | e-Health Record registry immunization notices |
|  | C-19 Resilience | NA |
|  | MAP-19 | Follow-up questionnaire (2^nd^-14^th^ January 2022) |
|  | MoBa | Biweekly follow-up questionnaires (2^nd^ February 2021-18^th^ February 2022) |
|  | CovidLife | Scotland National COVID -19 vaccination data (8^th^ December 2020 – 18^th^ February 2022) |
|  | Omtanke2020 | Annual follow-up questionnaire (1^st^ December 2021-18^th^ February 2022) |
| Covariates | | |
| Sociodemographic covariates (age, sex) | EstBB-C19 | Based on national personal identification number drawn at recruitment |
|  | EstBB-EHR | Based on national personal identification number drawn at recruitment |
|  | C-19 Resilience | Baseline questionnaire (24^th^ April 2020-29^th^ December 2020) |
|  | MAP-19 | Baseline questionnaire (31^st^ March 2020-7^th^ April 2020) |
|  | MoBa | First MoBa questionnaires filled out by females (mothers) and males (fathers) during pregnancy (recruitment 1999-2009) |
|  | CovidLife | Baseline questionnaire (COVIDLife1: 17th April -7th June 2020) |
|  | Omtanke2020 | Baseline questionnaire (9^th^ June 2020-26th December 2020) |
| Smoking status | EstBB-C19 | C-19 questionnaire (10^th^ May 2020 – 26^th^ December 2020) |
|  | EstBB-EHR | Baseline questionnaire completed upon joining EstBB (up to 26^th^ December 2020) |
|  | C-19 Resilience | Baseline questionnaire and monthly follow-up questionnaires (24^th^ April 2020-29^th^ December 2020) |
|  | MAP-19 | NA |
|  | MoBa | Biweekly COVID-19 follow-up questionnaires (10^th^ June 2020-23^rd^ June 2020) |
|  | CovidLife | Baseline questionnaire (COVIDLife1: 17th April -7th June 2020) |
|  | Omtanke2020 | Baseline questionnaire (9^th^ June 2020-26th December 2020) |
| Previous COVID-19 infection | EstBB-C19 | C-19 questionnaire (10^th^ May 2020 – 26^th^ December 2020) |
|  | EstBB-EHR | e-Health Record registry (up to 26^th^ December 2020) |
|  | C-19 Resilience | Baseline questionnaire and monthly follow-up questionnaires (24^th^ April 2020-29^th^ December 2020) |
|  | MAP-19 | Baseline questionnaire (31^st^ March 2020-7^th^ April 2020) |
|  | MoBa | Baseline COVID-19 questionnaire and biweekly COVID-19 follow-up questionnaires (31st March 2020-19^th^ December 2020) |
|  | CovidLife | Electronic Communication of Surveillance in Scotland (ECOSS) data on COVID-19 infections (29^th^ January 2020 – 8^th^ December 2020) |
|  | Omtanke2020 | Baseline and monthly follow-up questionnaires (9^th^ June 2020-26^th^ December 2020) |
| Physical comorbidity status | EstBB-C19 | C-19 questionnaire (10^th^ May 2020 – 26^th^ December 2020) |
|  | EstBB-EHR | Diagnosis from HIF bills (2004 - 26^th^ December 2020) |
|  | C-19 Resilience | Baseline questionnaire (24^th^ April 2020-29^th^ December 2020) |
|  | MAP-19 | NA |
|  | MoBa | For chronic renal failure and immunological conditions: biweekly COVID-19 follow-up questionnaires (5^th^-18^th^ December 2020)  For other conditions: baseline COVID-19 questionnaire (31^st^ March 2020-12^th^ May 2020) |
|  | CovidLife | Baseline questionnaire (17th April -7th June 2020) |
|  | Omtanke2020 | Baseline questionnaire (9^th^ June 2020-26th December 2020) |

NA: not applicable (data no available in cohort)

EstBB-C19 = The Estonian Biobank COVID-19 Cohort; EstBB-EHR = The Estonian Biobank electronic health records; C19-Resilience = The Icelandic COVID-19 National Resilience Cohort; MAP-19 = The Norwegian COVID-19, Mental Health and Adherence Project; MoBa = The Norwegian Mother, Father and Child Cohort Study

### **Supplementary Table 5:** ICD-10 codes used to define mental illness, and types of mental illness, in the Swedish register population.

| Mental Illness | ICD-10 codes |
| --- | --- |
| Any mental illness | F10-F19, F20-F29, F32, F33, F40, F41, F43 |
| Substance use disorder (not including tobacco and alcohol) | F11-F19 (excluding F17) |
| Alcohol use disorder | F10 |
| Tobacco use disorder | F17 |
| Psychotic disorders (schizophrenia and non-affective psychotic disorders) | F20-F29 |
| Depression | F32, F33 |
| Anxiety | F40, F41 |
| Stress-related disorders | F43 |

### **Supplementary Table 6:** ATC codes used to identify psychiatric medication in the Swedish register population.

| Psychiatric medication | ATC codes |
| --- | --- |
| Any psychiatric medication | N06A, N05B, N05C, N05A |
| Antidepressants | N06A |
| Anxiolytics | N05B |
| Hypnotics/sedatives | N05C |
| Antipsychotics | N05A |

### **Supplementary Table 7:** Distribution of sociodemographic variables in the total included COVIDMENT study population, and in each participating cohort, presented as N (%) or mean [SD].

| Variables | EstBB-C19 (Estonia) (n=5,633) | EstBB-EHR (Estonia) (n=183,332) | C-19 Resilience (Iceland) (n=10,417) | MAP-19 (Norway) (n=3,894) | MoBa (Norway) (n=102,811) | CovidLife (Scotland) (n=4,760) | Omtanke2020 (Sweden) (n=14,451) | Total (N=325,298) |
| --- | --- | --- | --- | --- | --- | --- | --- | --- |
| Sex | | | | | | | | |
| Female | 4,240 (75.3%) | 120,436 (65.7%) | 7,115 (68.3%) | 3,002 (77.1%) | 61,973 (60.3%) | 2,989 (62.8%) | 12,057 (83.4%) | 211,812 (65.1%) |
| Male | 1,393 (24.7%) | 62,896 (34.3%) | 3,193 (30.7%) | 883 (22.7%) | 40,838 (39.7%) | 1,767 (37.1%) | 2,394 (16.6%) | 113,364 (34.9%) |
| Other | 0 (0.0%) | 0 (0.0%) | 15 (0.1%) | 0 (0.0%) | 0 (0.0%) | 0 (0.0%) | 0 (0.0%) | 15 (0.0%) |
| Missing | 0 (0.0%) | 0 (0.0%) | 94 (0.9%) | 9 (0.2%) | 0 (0.0%) | 4 (0.1%) | 0 (0.0%) | 107 (0.0%) |
| Age group, years | | | | | | | | |
| 18-29 | 589 (10.5%) | 23,751 (13.0%) | 427 (4.1%) | 1,552 (39.9%) | 0 (0.0%) | 43 (0.9%) | 1,399 (9.7%) | 27,761 (8.5%) |
| 30-39 | 1,426 (25.3%) | 40,308 (22.0%) | 909 (8.7%) | 888 (22.8%) | 8,860 (8.6%) | 358 (7.5%) | 2,053 (14.2%) | 54,802 (16.9%) |
| 40-49 | 1,337 (23.7%) | 37,637 (20.5%) | 1,727 (16.6%) | 660 (16.9%) | 65,942 (64.2%) | 587 (12.3%) | 2,462 (17.0%) | 110,352 (33.9%) |
| 50-59 | 1,259 (22.4%) | 33,892 (18.5%) | 2,736 (26.2%) | 465 (11.9%) | 26,549 (25.8%) | 1,092 (22.9%) | 3,335 (23.1%) | 69,328 (21.3%) |
| 60-69 | 727 (12.9%) | 26,667 (14.5%) | 2,943 (28.3%) | 259 (6.7%) | 1,252 (1.2%) | 1,662 (34.9%) | 2,918 (20.2%) | 36,428 (11.2%) |
| 70 or above | 295 (5.2%) | 21,077 (11.5%) | 1,675 (16.1%) | 70 (1.8%) | 67 (0.1%) | 1,011 (21.3%) | 2,284 (15.8%) | 26,479 (8.1%) |
| Missing | 0 (0.0%) | 0 (0.0%) | 0 (0.0%) | 0 (0.0%) | 141 (0.1%) | 7 (0.2%) | 0 (0.0%) | 148 (0.1%) |
| Mean [SD] age, years | 46.3 [13.6] | 48.1 [16.2] | 56.1 [13.4] | 36.9 [13.8] | 46.5 [5.3] | 59.4 [12.1] | 52.3 [15.6] | 48.0 [2.9] |
| Previous COVID-19 infection | | | | | | | | |
| Yes | 912 (16.2%) | 3,772 (2.0%) | 327 (3.1%) | 5 (0.1%) | 2,007 (2.0%) | 83 (1.7%) | 634 (4.4%) | 7,740 (2.4%) |
| No | 4,721 (83.8%) | 67,946 (37.1%) | 10,090 (96.9%) | 3,889 (99.9%) | 100,804 (98.0%) | 4,677 (98.3%) | 13,817 (95.6%) | 205,944 (63.3%) |
| Missing | 0 (0.0%) | 111,614 (60.9%) | 0 (0.0%) | 0 (0.0%) | 0 (0.0%) | 0 (0.0%) | 0 (0.0%) | 111,614 (34.3%) |
| Smoking status | | | | | | | | |
| Yes | 773 (13.7%) | 36,035 (19.7%) | 1,486 (14.3%) | 0 (0.0%) | 20,297 (19.8%) | 257 (5.4%) | 1,932 (13.4%) | 60,780 (18.7%) |
| No | 4,542 (80.6%) | 130,074 (70.9%) | 8,918 (85.6%) | 0 (0.0%) | 81,967 (79.7%) | 4,414 (92.7%) | 12,496 (86.5%) | 242,411 (74.5%) |
| Missing | 318 (5.7%) | 17,223 (9.4%) | 13 (0.1%) | 3,894 (100.0%) | 547 (0.5%) | 89 (1.9%) | 23 (0.1%) | 22,107 (6.8%) |
| Chronic physical conditions | | | | | | | | |
| 0 | 2,574 (45.7%) | 68,730 (37.5%) | 5,735 (55.1%) | 0 (0.0%) | 81879 (79.6%) | 3,546 (74.5%) | 9,250 (64.0%) | 171,714 (52.8%) |
| 1 | 1,805 (32.0%) | 55,229 (30.1%) | 3,099 (29.7%) | 0 (0.0%) | 17169 (16.7%) | 972 (20.4%) | 3,353 (23.2%) | 81,627 (25.1%) |
| ≥2 | 775 (13.8%) | 59,373 (32.4%) | 1,485 (14.3%) | 0 (0.0%) | 3763 (3.7%) | 217 (4.6%) | 1,218 (8.4%) | 66,831 (20.5%) |
| Missing | 479 (8.5%) | 0 (0.0%) | 98 (0.9%) | 3,894 (100.0%) | 0 (0.0%) | 25 (0.5%) | 630 (4.4%) | 5,126 (1.6%) |

EstBB-C19 = The Estonian Biobank COVID-19 Cohort; EstBB-EHR = The Estonian Biobank electronic health records; C19-Resilience = The Icelandic COVID-19 National Resilience Cohort; MAP-19 = The Norwegian COVID-19, Mental Health and Adherence Project; MoBa = The Norwegian Mother, Father and Child Cohort Study

### **Supplementary Table 8:** Uptake of COVID-19 vaccination in each included COVIDMENT cohort, overall and by presence of any mental illness diagnosis, presented as N (%).

|  | Uptake of first dose of a COVID-19 vaccine by 30^th^ September 2021 | | | Uptake of first dose of a COVID-19 vaccine by 18^th^ February 2022 | | | Uptake of second dose of a COVID-19 vaccine by 18^th^ February 2022 | | |
| --- | --- | --- | --- | --- | --- | --- | --- | --- | --- |
|  | Yes | No | Total | Yes | No | Total | Yes | No | Total |
| EstBB-C19 (Estonia) | 5,108 (90.7%) | 525 (9.3%) | 5,633 (100.0%) | 5,243 (93.1%) | 390 (6.9%) | 5,633 (100.0%) | 4,950 (94.4%) | 294 (5.6%) | 5,244 (100.0%) |
| Mental illness | 2,796 (90.0%) | 311 (10.0%) | 3,107 (100.0%) | 2,870 (92.4%) | 237 (7.6%) | 3,107 (100.0%) | 2,708 (94.4%) | 162 (5.6%) | 2,870 (100.0%) |
| No mental illness | 2,312 (91.5%) | 214 (8.5%) | 2,526 (100.0%) | 2,373 (93.9%) | 153 (6.1%) | 2,526 (100.0%) | 2,242 (94.4%) | 132 (5.6%) | 2,374 (100.0%) |
| Missing | 0 (0.0%) | 0 (0.0%) | 0 (0.0%) | 0 (0.0%) | 0 (0.0%) | 0 (0.0%) | 0 (0.0%) | 0 (0.0%) | 0 (0.0%) |
| EstBB-EHR (Estonia) | 150,387 (82.0%) | 32,945 (18.0%) | 183,332 (100.0%) | 157,983 (86.2%) | 25,349 (13.8%) | 183,332 (100.0%) | 143,877 (94.4%) | 8,469 (5.6%) | 152,346 (100.0%) |
| Mental illness | 76,855 (80.7%) | 18,353 (19.3%) | 95,208 (100.0%) | 81,038 (85.1%) | 14,170 (14.9%) | 95,208 (100.0%) | 73,634 (94.2%) | 4,541 (5.8%) | 78,175 (100.0%) |
| No mental illness | 73,532 (83.4%) | 14,592 (16.6%) | 88,124 (100.0%) | 76,945 (87.3%) | 11,179 (12.7%) | 88,124 (100.0%) | 70,243 (94.7%) | 3,928 (5.3%) | 74,171 (100.0%) |
| Missing | 0 (0.0%) | 0 (0.0%) | 0 (0.0%) | 0 (0.0%) | 0 (0.0%) | 0 (0.0%) | 0 (0.0%) | 0 (0.0%) | 0 (0.0%) |
| C-19 Resilience (Iceland) | 9,117 (87.5%) | 1,300 (12.5%) | 10,417 (100.0%) | NA | NA | NA | NA | NA | NA |
| Mental illness | 2,460 (85.0%) | 435 (15.0%) | 2,895 (100.0%) | NA | NA | NA | NA | NA | NA |
| No mental illness | 6,446 (88.5%) | 837 (11.5%) | 7,283 (100.0%) | NA | NA | NA | NA | NA | NA |
| Missing | 211 (88.3%) | 28 (11.7%) | 239 (100.0%) | NA | NA | NA | NA | NA | NA |
| MAP-19 (Norway) | 2,529 (78.3%) | 702 (21.7%) | 3,231 (100.0%) | 2,523 (97.2%) | 74 (2.8%) | 2,597 (100.0%) | 2,482 (98.4%) | 41 (1.6%) | 2,523 (100.0%) |
| Mental illness | 425 (73.9%) | 150 (26.1%) | 575 (100.0%) | 503 (95.8%) | 22 (4.2%) | 525 (100.0%) | 492 (97.8%) | 11 (2.2%) | 503 (100.0%) |
| No mental illness | 1,736 (79.6%) | 446 (20.4%) | 2,182 (100.0%) | 2,020 (97.5%) | 52 (2.5%) | 2,072 (100.0%) | 1,990 (98.5%) | 30 (1.5%) | 2,020 (100.0%) |
| Missing | 368 (77.6%) | 106 (22.4%) | 474 (100.0%) | 0 (0.0%) | 0 (0.0%) | 0 (0.0%) | 0 (0.0%) | 0 (0.0%) | 0 (0.0%) |
| MoBa (Norway) | 91,424 (89.5%) | 10,704 (10.5%) | 102,128 (100.0%) | 94,500 (91.9%) | 8,311 (8.1%) | 102,811 (100.0%) | 91,442 (96.8%) | 3,058 (3.2%) | 94,500 (100.0%) |
| Mental illness | 13,688 (89.0%) | 1,696 (11.0%) | 15,384 (100.0%) | 14,163 (91.4%) | 1,333 (8.6%) | 15,496 (100.0%) | 13,658 (96.4%) | 505 (3.6%) | 14,163 (100.0%) |
| No mental illness | 77,736 (89.6%) | 9,008 (10.4%) | 86,744 (100.0%) | 80,337 (92.0%) | 6,978 (8.0%) | 87,315 (100.0%) | 77,784 (96.8%) | 2,553 (3.2%) | 80,337 (100.0%) |
| Missing | 0 (0.0%) | 0 (0.0%) | 0 (0.0%) | 0 (0.0%) | 0 (0.0%) | 0 (0.0%) | 0 (0.0%) | 0 (0.0%) | 0 (0.0%) |
| CovidLife (Scotland) | 4,477 (94.1%) | 283 (5.9%) | 4,760 (100.0%) | 4,484 (94.2%) | 276 (5.8%) | 4,760 (100.0%) | 4,461 (100.0%) | 1 (0.0%) | 4,462 (100.0%) |
| Mental illness | 1,151 (93.6%) | 79 (6.4%) | 1,230 (100.0%) | 1,151 (93.6%) | 79 (6.4%) | 1,230 (100.0%) | 1,145 (100.0%) | 0 (0.0%) | 1,145 (100.0%) |
| No mental illness | 3,296 (94.2%) | 203 (5.8%) | 3,499 (100.0%) | 3,303 (94.4%) | 196 (5.6%) | 3,499 (100.0%) | 3,290 (100.0%) | 1 (0.0%) | 3,291 (100.0%) |
| Missing | 30 (96.8%) | 1 (3.2%) | 31 (100.0%) | 30 (96.8%) | 1 (3.2%) | 31 (100.0%) | 26 (100.0%) | 0 (0.0%) | 26 (100.0%) |
| Omtanke2020 (Sweden) | 4,939 (92.7%) | 387 (7.3%) | 5,326 (100.0%) | 14,154 (97.9%) | 297 (2.1%) | 14,451 (100.0%) | 5,227 (98.1%) | 102 (1.9%) | 5,329 (100.0%) |
| Mental illness | 1,666 (91.9%) | 147 (8.1%) | 1,813 (100.0%) | 4,230 (97.4%) | 112 (2.6%) | 4,342 (100.0%) | 1,783 (98.2%) | 32 (1.8%) | 1,815 (100.0%) |
| No mental illness | 3,116 (93.1%) | 232 (6.9%) | 3,348 (100.0%) | 9,634 (98.3%) | 170 (1.7%) | 9,804 (100.0%) | 3,281 (98.0%) | 68 (2.0%) | 3,349 (100.0%) |
| Missing | 157 (95.2%) | 8 (4.8%) | 165 (100.0%) | 290 (95.1%) | 15 (4.9%) | 305 (100.0%) | 163 (98.8%) | 2 (1.2%) | 165 (100.0%) |

### **Supplementary Table 9:** Meta-analyses heterogeneity measure results in the included COVIDMENT study population.

|  | Heterogenity (I^2^) | | |
| --- | --- | --- | --- |
| Variable | Any mental illness diagnosis | Anxiety symptoms | Depressive symptoms |
| First dose of a COVID-19 vaccine by 30^th^ September 2021 | 91.4%*** | 60.7%* | 63.0%** |
| First dose of a COVID-19 vaccine by 18^th^ February 2022 | 80.0%*** | 73.7%** | 73.4%** |
| Second dose of a COVID-19 vaccine by 18^th^ February 2022 | 60.7% | 2.0% | 53.4% |

***p<0.001; **p<0.01; *p<0.05

### **Supplementary Table 10:** Sensitivity model results (pooled prevalence ratio [PR] (95% CI)), according to the presence of any mental illness diagnosis, in the included COVIDMENT study population.

|  | Sensitivity analysis 1 | | Sensitivity analysis 2 | |
| --- | --- | --- | --- | --- |
|  | Pooled PR (95% CI) | I^2^ | Pooled PR (95% CI) | I^2^ |
| Outcome | | |  |  |
| First dose of a COVID-19 vaccine by 30^th^ September 2021 | 0.99 (0.96-1.01) | 91.3%** | 0.99 (0.98-1.00) | 87.1%*** |
| First dose of a COVID-19 vaccine by 18^th^ February 2022 | 0.99 (0.99-1.00) | 18.2% | 0.99 (0.98-1.00) | 76.8%* |
| Second dose of a COVID-19 vaccine by 18^th^ February 2022 | NA | NA | 1.00 (0.99-1.00) | 22.5% |

Sensitivity analysis 1: exclusion of cohorts which used electronic health records for the definition of exposure and/or outcome variables

Sensitivity analysis 2: exclusion of individuals with any chronic physical conditions

NA: not applicable (sensitivity analysis 1 was not run for the ‘second dose of a COVID-19 vaccine by 18^th^ February 2022’ as only two cohort could be included)

***p<0.001; **p<0.01; *p<0.05

### **Supplementary Table 11:** Uptake of COVID-19 vaccination by type of mental illness diagnosis and prescribed psychiatric medication use, in the included Swedish register population, presented as N (%).

|  | First dose of a COVID-19 vaccine by 30^th^ September 2021 | | | Second dose of a COVID-19 vaccine by 30^th^ November 2021 | | |
| --- | --- | --- | --- | --- | --- | --- |
|  | Yes | No | Total | Yes | No | Total |
| Type of mental illness | | | | | | |
| Substance use disorder* | 96,666 (73.9%) | 34,122 (26.1%) | 130,788 (100.0%) | 91,661 (94.8%) | 5,005 (5.2%) | 96,666 (100.0%) |
| Alcohol use disorder | 116,345 (79.7%) | 29,664 (20.3%) | 146,009 (100.0%) | 111,715 (96.0%) | 4,630 (4.0%) | 116,345 (100.0%) |
| Tobacco use disorder | 81,827 (81.2%) | 18,896 (18.8%) | 100,723 (100.0%) | 78,968 (96.5%) | 2,859 (3.5%) | 81,827 (100.0%) |
| Psychotic disorders | 99,597 (78.9%) | 26,686 (21.1%) | 126,283 (100.0%) | 96,067 (96.5%) | 3,530 (3.5%) | 99,597 (100.0%) |
| Depression | 178,925 (80.7%) | 42,697 (19.3%) | 221,622 (100.0%) | 172,252 (96.3%) | 6,672 (3.7%) | 178,924 (100.0%) |
| Anxiety | 193,133 (78.7%) | 52,412 (21.4%) | 245,545 (100.0%) | 185,299 (95.9%) | 7,834 (4.1%) | 193,133 (100.0%) |
| Stress-related disorders | 141,489 (78.7%) | 38,344 (21.3%) | 179,833 (100.0%) | 136,120 (96.2%) | 5,369 (3.8%) | 141,489 (100.0%) |
| Type of psychiatric medication | | | | | | |
| Antidepressants | 1,127,619 (87.7%) | 158,523 (12.3%) | 1,286,142 (100.0%) | 1,104,282 (97.9%) | 23,336 (2.1%) | 1,127,618 (100.0%) |
| Anxiolytics | 673,869 (85.9%) | 110,972 (14.1%) | 784,841 (100.0%) | 658,987 (97.8%) | 14,881 (2.2%) | 673,868 (100.0%) |
| Hypnotics/sedatives | 940,546 (88.3%) | 125,222 (11.8%) | 1,065,768 (100.0%) | 922,633 (98.1%) | 17,911 (1.9%) | 940,544 (100.0%) |
| Antipsychotics | 157,254 (81.3%) | 36,252 (18.7%) | 193,506 (100.0%) | 152,536 (97.0%) | 4,718 (3.0%) | 157,254 (100.0%) |

*Not including alcohol and tobacco use disorders.

### **Supplementary Table 12:** Stratified model results (prevalence ratio (95% CI)) performed in the Swedish register population, using ‘any mental illness’ and ‘any medication’ as exposure variables, and ‘first dose of COVID-19 vaccination by 30^th^ September 2021’ and ‘second dose of a COVID-19 vaccine by 30^th^ November 2021’ as outcome variables.

|  | Prevalence Ratio (95% CI) | | | |
| --- | --- | --- | --- | --- |
|  | First dose of a COVID-19 vaccine by 30^th^ September 2021 | | Second dose of a COVID-19 vaccine by 30^th^ November 2021 | |
|  | Any mental illness | Any medication | Any mental illness | Any medication |
| Sex | | | | |
| Male | 0.97 (0.97-0.97)*** | 1.02 (1.02-1.02)*** | 0.99 (0.98-0.99)*** | 1.00 (1.00-1.00)*** |
| Female | 1.00 (0.99-1.00)*** | 1.03 (1.03-1.03)*** | 0.99 (0.99-0.99)*** | 1.00 (1.00-1.00)*** |
| Chronic physical condition | | | | |
| 0 | 0.98 (0.98-0.99)*** | 1.03 (1.03-1.03)*** | 0.99 (0.99-0.99)*** | 1.00 (1.00-1.00)*** |
| ≥1 | 0.97 (0.97-0.97)*** | 1.01 (1.01-1.01)*** | 0.99 (0.99-0.99)*** | 1.00 (1.00-1.00)*** |

***p<0.001; **p<0.01; *p<0.05

### **Supplementary Figure 1:** Flow chart of COVIDMENT study population.

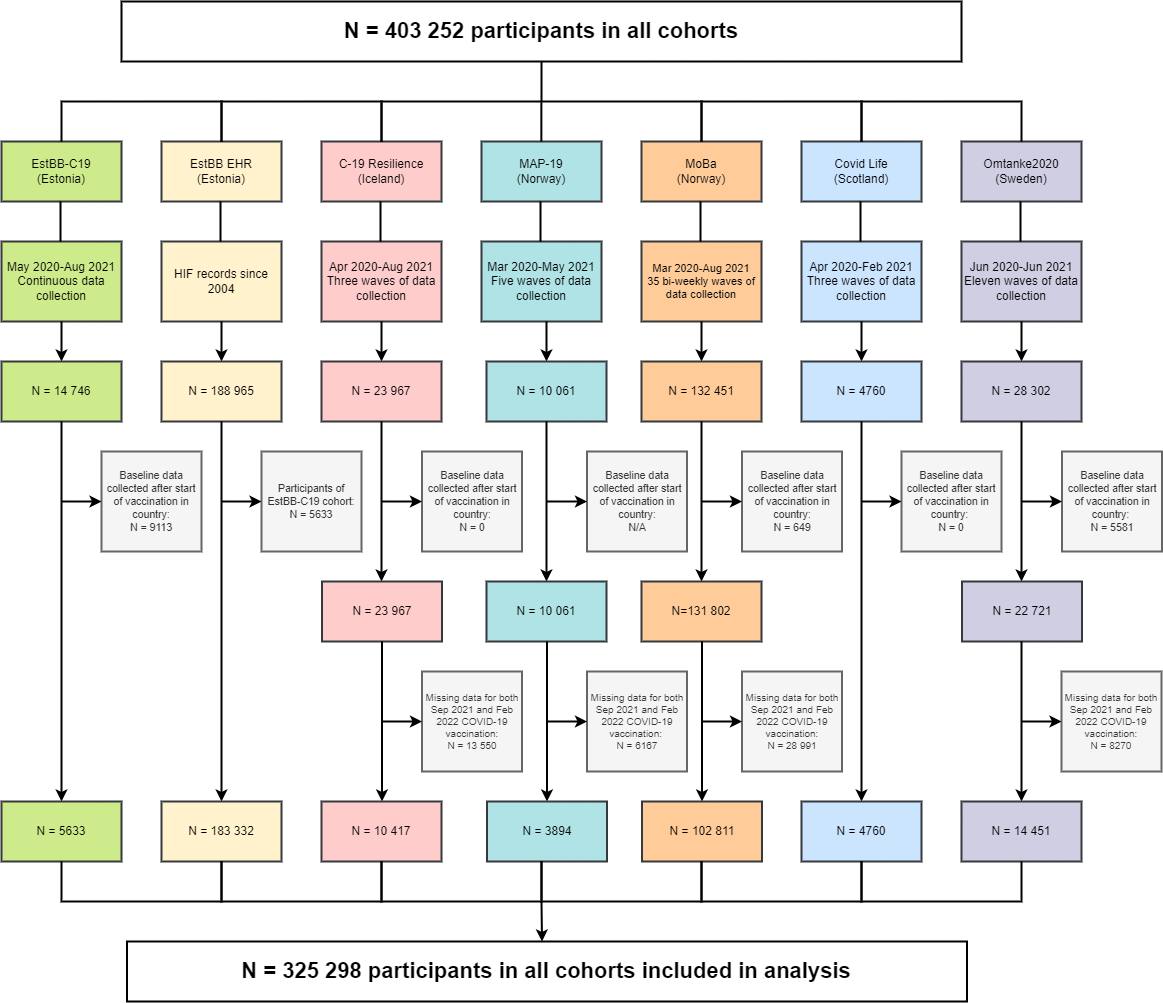

*CovidLife (Scotland): only CovidLife participants with linked vaccination data from Generation Scotland were included in the study population.
